# An implementation-effectiveness trial investigating feasibility, safety and efficacy of Best pRacticE guideline cAre physioTHErapy with addition of protocolised ventilator hyperinflation on patient-important outcomes for critically ill adults intUbated and ventilated with Pneumonia. (BREATHE UP)

**DOI:** 10.1101/2024.11.10.24317069

**Authors:** Lisa van der Lee, Adrian Regli, Christopher Allen, Matthew Anstey, Robert Blakeman, Linda Denehy, Diane Dennis, Mercedes Elliott, Anne-Marie Hill, Kwok-Ming Ho, Wendy Jacob, Angela Jacques M, Lisa Marsh, Mark Palermo, Bradley Wibrow, George Ntoumenopoulos

## Abstract

Pneumonia is a common cause for intensive care unit (ICU) admission for breathing support from a mechanical ventilator, resulting in high morbidity, mortality, and healthcare costs. Physiotherapists working in ICU treat patients with critical pneumonia using a range of respiratory treatments to improve breathing, hasten weaning from the ventilator, and restore functional independence. Evidence supports the short-term benefits of these treatments, however currently no standard of physiotherapy practice exists for these ICU patients. Despite a plethora of studies which have shown that physiotherapy treatments, such as ventilator lung hyperinflation (VHI), aid the lungs to work better short-term, there are no studies to date which investigate the effect of these important treatments on meaningful patient-important outcomes, such as earlier weaning from the ventilator, earlier hospital discharge, less breathlessness and lung secretions, short- and longer-term disability, health-related quality of life and survival. Through previous multiphase mixed-methods research, a clinical practice guideline was developed outlining best physiotherapy practice for this ICU patient cohort. This proposed research involves conducting a multicentre hybrid implementation-effectiveness randomised, controlled trial which aims to: evaluate implementation of physiotherapy guideline care for management of pneumonia in ICU regarding feasibility, safety and acceptability; and to determine the safety and effectiveness of a standardised dosage of daily ventilator hyperinflation in addition to standard guideline care on patient-important outcomes, health system outcomes; and cost effectiveness. This study will address critical knowledge gaps in the ICU physiotherapy evidence base regarding the impact of a course of standardised, best practice respiratory physiotherapy intervention during the ICU stay for patients with critical pneumonia requiring invasive mechanical ventilation, provide valuable information for clinicians to inform clinical practice regarding treatable traits and streamlining care to promote clinical efficiency, and determine the impact of respiratory physiotherapy care on patient-important and health service outcomes.

**Trial registration:** ANZ Clinical trials Registry number ACTRN12624001130550.

## INTRODUCTION

Pneumonia was a leading cause of death worldwide pre-pandemic [1, 2], with 1.23 million deaths in people aged over 70 years, and over 1.6 million aged over 50 years [1]. This is significant in the context of a global ageing population, with a population aged over 60 years expected to double to 2.1 billion by 2050 [3]. In Australia, pneumonia due to *Streptococcus pneumoniae* is a leading cause of hospitalisation, costing over AUD$ 55 million [4], with 10- 30% of people hospitalised requiring ICU admission for breathing support by a mechanical ventilator [4, 5], with a median length of stay in ICU of 10 days [6]. However, at a cost of AUD $ 4875 per ICU stay day, the provision of intensive care is very expensive [7], highlighting the significant clinical and economic burden of pneumonia in Australia.

Physiotherapists working in ICU treat patients with severe pneumonia [8-11], to improve clearance of lung secretions (sputum) and ease breathing, facilitate weaning from the mechanical ventilator and restore functional independence [12, 13]. Facilitation of early weaning from the ventilator is critical, to minimise the occurrence of hospital-acquired infections, and sequelae of bed rest, such as pressure injuries, joint contractures, and ICU acquired-weakness, all of which increase care costs, ICU and hospital length of stay, and increase risk of long-term disability [14]. Evidence supports the short-term benefits of these treatments, however currently no standard of physiotherapy practice or protocols exist for these frequently admitted ICU patients and evidence for the effect of these physiotherapy treatments on patient-centred outcomes is lacking [15]. Further, physiotherapy clinical practice for this ICU cohort is variable [10, 16] and highly dependent on the clinical decisions of often junior and inexperienced physiotherapists. Variability in clinical practice primarily stems from a lack of high-quality evidence to guide practice, which has the potential to result in unfavourable outcomes for these critically ill patients [8, 9].

Despite a plethora of studies that have shown that respiratory physiotherapy treatments, such as ventilator hyperinflation techniques, *do aid the lungs to work better in the short-term* [15, 17-24], there are no studies to date that investigate the effect of these important treatments on meaningful patient-centred outcomes, such as earlier weaning from the ventilator, less symptoms of sputum and breathlessness, earlier hospital discharge, less short- and long-term disability, and improved quality of life, and survival [15]. Physiotherapy treatments commenced early when the patient is on the ventilator, to improve the patients’ ability to breathe by themselves sooner and rehabilitate earlier to regain their function and independence, have the potential to reduce time spent in ICU, hasten discharge from hospital and facilitate return to independent living in the community.

A world-first international clinical practice guideline for best physiotherapy management of adult patients requiring invasive mechanical ventilation in ICU for community-acquired pneumonia [25] was developed through a mixed-methods doctoral research program, which included: a systematic review and meta-analysis [15], which demonstrated significant short-term benefits from respiratory treatments that temporarily increase the patient’s breath volume when they are on the mechanical ventilator (termed ventilator hyperinflation - VHI), which clears sputum and aids the lungs to expand easier; a national survey of physiotherapy clinical practice [10, 16]; development of international expert consensus [11], locally validated by senior multidisciplinary ICU clinicians for validity and applicability in clinical practice [26]; and guideline development according to JBI [27] and GRADE [28] methodology. This guideline comprises 26 recommendations, encompassing physiotherapy assessment, patient prioritisation, and treatment including humidification, patient positioning, lung hyperinflation techniques (such as VHI), manual chest wall techniques, normal saline instillation and active treatment and mobilisation [25]. Further research will investigate patient and family preferences for treatment, to enable the incorporation of consumer perspectives into the guideline [27, 29].

Before widespread clinical use, the next stage of research involves implementation and evaluation of the guideline in clinical practice. To maximise clinical utility, the guideline will be implemented and evaluated for patients with both community-acquired pneumonia (CAP) and hospital-acquired pneumonia (HAP). An important domain in the guideline of lung hyperinflation was not able to be standardised as an intervention technique due to a lack of research. However, recent research by Jacob et al [23] found in a randomised crossover trial that a more sophisticated VHI lung hyperinflation protocol delivered using the mechanical ventilator that achieved a calculated target increase in lung breath volume, personalised for each patient according to their individual calculated inspiratory reserve volume but limited by peak pressure, improved sputum clearance better than the standard methods previously used [21]. The impact a course of physiotherapy VHI treatment has on patient outcomes, or what dosage of treatment is best is unknown.

This proposed world-first research will provide vital evidence to determine whether a course of protocolised physiotherapy ventilator lung hyperinflation treatment, added to guideline care, is feasible, safe, cost-effective and improves patient-important outcomes of disability, ventilator-free days, 90-day mortality, time spent in hospital, symptom severity and health-related quality of life for ICU patients with pneumonia requiring mechanical ventilation.

Therefore, the objectives of this research are:

1. To determine the feasibility (acceptability and fidelity) of implementing the physiotherapy guideline in patients with pneumonia, incorporating the new VHI treatment by Jacob et al [23].
2. To explore pneumonia phenotypes which demonstrate treatable traits that are amenable to respiratory physiotherapy intervention commenced during the acute period of invasive mechanical ventilation.
3. To determine the effect of a course of new VHI treatment method [23], at a set dosage and frequency of three times daily for the duration of the period that patients are on the mechanical ventilator on symptomatology and patient-centred outcomes.
4. To determine the cost-effectiveness of physiotherapy guideline care incorporating a course of new VHI treatment for pneumonia requiring invasive ventilation in ICU.

## METHODS

### Design

This is a two-arm, multicentre, comparative, assessor blinded, randomised, controlled, hybrid (Type 1 examining both efficacy and feasibility) superiority trial [30, 31]. This trial is designed according to the SPIRIT guidelines [32], will be conducted in accordance with the principles of Good Clinical Practice [33] and reported according to the CONSORT [34] and the complimentary StaRI standards for implementation studies [35]. Eligible patients will be randomised to receive either guideline physiotherapy care, or guideline physiotherapy care incorporating protocolised VHI [23] at a set daily dosage.

#### Consumer and Community Involvement

A preceding consumer co-designed qualitative research study has been conducted to inform this trial, whereby ICU survivors of critical pneumonia requiring invasive mechanical ventilation, and a close family member, were interviewed to explore their ICU experience with respiratory physiotherapy and to determine their values and preferences for treatment, for incorporation into the physiotherapy guideline, prior to evaluation in practice. Consumer consultation for co-design has and will continue to occur throughout the research program, provided by a consumer representative sourced through the Consumer and Community Involvement Program, who has many years of experience with chronic pneumonia frequently requiring admissions to ICU for treatment including mechanical ventilation. This consumer provided input into the protocol and the outcome measures for the trial.

#### Ethical considerations for the trial

Ethics approval has been granted from South Metropolitan Health Service Human Research and Ethics Committee in Western Australia (RGS 6734) and reciprocal ethics approvals and governance approvals will be sought at each participating site.

Currently, clinical equipoise exists pertaining to the efficacy of respiratory physiotherapy treatments commonly provided in ICU for mechanically ventilated patients [15, 36] and there is evidence of significant variability in delivery of physiotherapy treatment mode, technique, duration and frequency in clinical practice [10, 16], both within and between health service providers. Patients enrolled in this study will be critically ill and likely sedated and hence unable to make sufficient judgements to provide their own informed consent for study participation. Consent for participation in this study will be sought from a substitute decision maker for a research candidate according to local jurisdiction legislative requirements [37], by research trained staff. Following enrolment in the study via the surrogate decision maker, written informed consent for ongoing consent to continue as a research participant will be sought from the participant once they have capacity to make reasonable judgements regarding their participation in the research. The two arms of this study represent treatment options which are both considered under the umbrella of current practice, with the physiotherapy guideline ensuring that all patients enrolled under the study will receive minimum best practice physiotherapy care. Permission to obtain contact details from the surrogate decision maker for 90-day follow-up contact will also be sought by research trained staff at the sites, in case the patient is unable to be contacted for any reason. If a patient has subsequently died, all data collected will still be used in the analysis according to intention to treat principle, unless consent to use data has been withdrawn prior to analysis.

### Participants and setting

This study will be conducted in the Intensive Care Unit (ICU) setting, in tertiary hospitals in Australia. Patients admitted to ICU for treatment of respiratory failure will be screened for eligibility by trained ICU staff. Eligible patients are those who fulfill all inclusion criteria without any exclusion criteria.

#### Inclusion Criteria

- ICU admission with diagnosis of pneumonia made by a medical practitioner.
- Aged ≥ 18 years
- Intubated and mechanically ventilated, expected to remain so for > 24 hours, not planned for extubation this calendar day.
- Evidence of consolidation or volume loss on CXR, CT scan or lung ultrasound.
- Evidence of sputum on clinical assessment by a physiotherapist, e.g. auscultation, palpation, endotracheal suction, waveform analysis.

#### Exclusion criteria

- Death imminent or the treating clinician believes that death during this hospital admission is inevitable.
- Treating clinician believes that trial participation is not in the best interests of the patient.
- Receiving invasive mechanical ventilation for > 5 days
- Peak airway pressure > 35cmH20 consistently for ≥ 2 hours
- Progression to severe ARDS according to Berlin Criteria [38]
- Requirement for extracorporeal membrane oxygenation
- Severe bronchospasm
- Undrained pneumothorax or bronchopleural fistula
- Pulmonary haemorrhage
- Lung transplantation or recent lung surgery with bronchial resection
- Unable to communicate in English
- Pregnancy
- Underlying neurological or myopathic condition
- Documented cognitive impairment
- BMI≥35
- Hospitalisation for > 7 days prior to ICU
- Presence of active cancer or active use of chemotherapeutic agents or neutropenia
- Unlikely to be available for 3-month follow-up (resides overseas)

### Interventions

#### Study intervention

Patients will be randomised to receive either guideline physiotherapy care incorporating varied treatment modes which may include manual hyperinflation using a manual resuscitation bag or conservative VHI using the ventilator, delivered at a variable daily dosage according to the discretion of the treating physiotherapist (control arm), or guideline physiotherapy care incorporating the new VHI treatment [23] at a set daily dosage and frequency of 3 times per day (treatment arm), as outlined below. Figure 1 indicates patient flow through the study.

**Figure 1:**
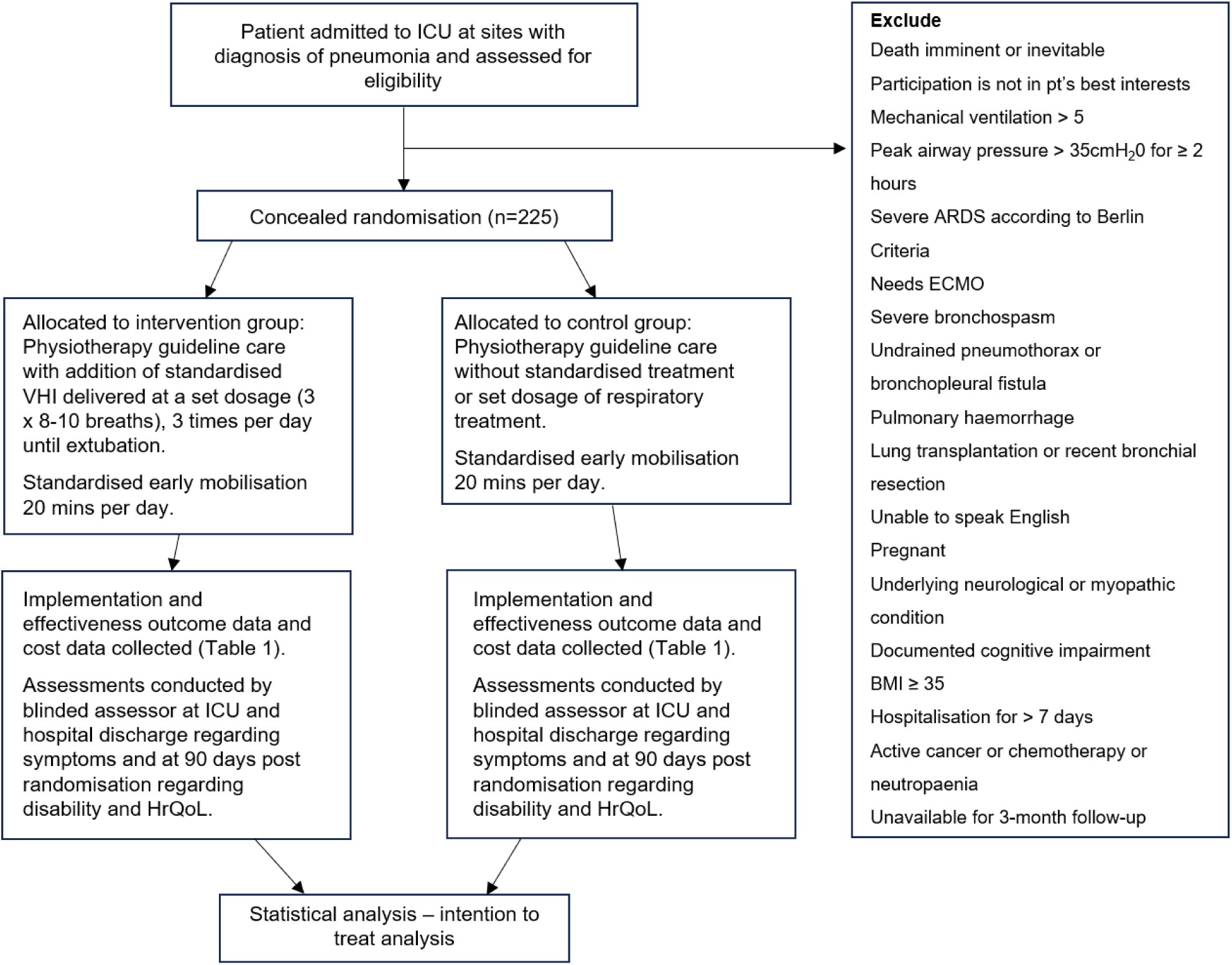
Study consort diagram

#### Comparator

Physiotherapy guideline care [25] will be provided without the new VHI treatment. For this arm of the trial, the physiotherapists will be provided with a guideline prompt tool (Appendix 1) but will not be provided with additional guidance regarding the mode, duration and frequency of the intervention(s) to provide for respiratory physiotherapy treatment. As per standard care, treatment will be provided based on local policies, with the mode and dosage of treatment being delivered at the discretion of the treating physiotherapist, based on their clinical judgement.

All aspects of management other than physiotherapy treatment will occur according to individual unit practice with no restrictions placed on concomitant care.

#### Sequence generation

A permuted block randomisation method with variable block sizes, stratified by site will be used to allocate participants to either standard guideline care or intervention group in a 1:1 ratio. Computerised randomisation and allocation concealment will be performed by research trained staff at sites using REDCap – Research Electronic Data Capture, a secure, web-based research data management application. Research staff will notify the intensive care physiotherapists of the group allocation following randomisation.

#### Blinding

It is not feasible to blind treatment providers in the ICU; but this is common to all process of care studies conducted in ICU, including successful, large RCTs. However, clinicians who care for the patients following discharge from the ICU will not be explicitly informed about allocation. Patients will be blinded to which treatment arm they are allocated to, as they will be unaware of the treatments and frequency they receive during the period of mechanical ventilation whilst they are sedated. Assessment of respiratory symptoms at ICU and hospital discharge will be performed by staff unaware of treatment allocation, and clinicians who care for the patients following discharge from the ICU will not be explicitly informed about allocation.

#### Safety

It has been well established in the literature that physiotherapy treatments delivered to critically ill patients in ICU are safe [39]. This includes commonly used respiratory physiotherapy treatments such as lung hyperinflation techniques [17-22, 24, 36, 40, 41]. The protocolised VHI treatment technique by Jacob et al [23] to be used in this study was found to be both safe and effective in improving lung mechanics and clearing sputum in a crossover trial of 48 patients. Physiotherapists working in the ICU setting are highly skilled and trained healthcare professionals who work collaboratively as part of a multidisciplinary team [42].

All critically ill patients in ICU receiving mechanical ventilation are cared for by a dedicated bedside nurse as standard care. All physiotherapy interventions delivered in ICU heed well documented contraindications and precautions [13, 43], in line with hospital policies, to ensure optimal safety of patients and minimise occurrence of adverse events.

In the event of untoward significant physiological perturbation (based on local policies) during the respiratory physiotherapy intervention, including protocolised VHI treatment, which cannot be immediately corrected, the consultant or registrar must assess the patient and document if treatment can continue. A treatment session must be ceased if any of the following occur and cannot be immediately corrected:

- SpO2 < 85% for > 2 minutes during physiotherapy
- Ventilator dyssynchrony

Significant patient agitation (Ramsay Agitation-Sedation Scale (RASS) ≥ 2)

- Symptomatic BP drop MAP < 60, or 5mmHg below the target set by the treating intensivist, for greater than 2 minutes or increase in noradrenaline requirement by 0.15mcg/kg
- Haemodynamically significant rhythm change that does not constitute a serious adverse event (SAE) (e.g. bradycardia, ventricular ectopy or rapid atrial fibrillation).

Patients will be excluded from receiving physiotherapy treatment when vasoactive medications are ≥0.2mcg/kg/min [44].

#### Data collection and procedure

Data will be collected and analysed using the intention-to-treat principle. All data will be collected by trained staff at each participating site. A CRF worksheet may be used to collect the data. Data will be collected on a prespecified eCRF using REDCap, and supported by a study-specific data dictionary and including checks of logical consistency and automatic query generation. Daily ICU clinical data will be captured from existing ICU Clinical Information Systems or databases at each site and collected by research staff. Physiotherapy specific data pertaining to delivered episodes of care and treatment parameters of care delivered will be recorded at the point-of-care by the treating physiotherapist, on a CRF worksheet for later entry into REDCap by research trained staff. Data quality and protocol standardisation will be optimised by arranging a start-up meeting, and the CPI will meet regularly, either in-person or online, with the PIs from other sites to provide regular monitoring, guidance, and support.

Randomised patients will be followed up to death or 90 days post-randomisation whichever occurs first. Data will be collected for all randomised patients including those excluded at any stage. If consent for participation is withdrawn (or if the patient or family wish to opt-out), data will not be used unless consent to do so is obtained. Patients (or a proxy – generally a close family member) who are alive at 90 days after randomisation will be interviewed by the site research coordinator by telephone. Next of kin contact details will be collected for follow-up purposes in case the patient is not contactable. The research coordinator will administer the WHODAS 2.0 and EQ5D-5L questionnaires. Data for all patients will be analysed on an intention-to-treat principle. Table 1 outlines the data to be collected at each time point.

**Table 1:** Data collection and timepoints.

| Baseline | ICU Daily until Day 28 or ICU discharge if sooner | ICU discharge | Hospital discharge | 90 days |
| --- | --- | --- | --- | --- |
| • ICU admission date<br>• Age<br>• Sex assigned at birth<br>• Weight and height<br>• Usual residence<br>• Employment status<br>• ICU admission diagnosis<br>• Co-morbidities<br>• APACHE II score<br>• SOFA sepsis score<br>• Receipt of antibiotics<br>• Receipt of vasopressors<br>• Microbiology results<br>• Inflammatory markers (*WBC count and CRP) | • Microbiology results<br>• Sputum MC+S results<br>• Receipt of mechanical ventilation<br>• Receipt of vasopressors<br>• Receipt of inhaled nitric oxide<br>• Receipt of antibiotics<br>• Receipt of corticosteroids<br>• Physiotherapy data<br>• Timing and duration of treatment<br>• Guideline care provided (checklist)<br>• Barriers to treatment<br>• Sputum load and characteristics<br>• Ventilation settings for VHI<br>• Calculated and achieved VHI lung volumes and airway pressures.<br>• Inflammatory markers* (day 7)<br>• Respiratory rescue therapies (re-intubation, bronchoscopy, ECMO, prone, APRV, unplanned physiotherapy treatment)<br>• Adverse events and serious adverse events | • ICU discharge date<br>• ICU mortality<br>• Symptoms of breathlessness – at rest and on exertion (Edmonton VAS & Modified MRC Dyspnoea Scale)<br>• Sputum burden (VAS) | • ICU readmission<br>• Surgeries received<br>• Hospital mortality<br>• Hospital discharge date<br>• Hospital discharge destination<br>• Symptoms of breathlessness<br>• at rest and on exertion (Edmonton VAS & Modified MRC Dyspnoea Scale)<br>• Sputum burden (VAS) | • Disability (WHODAS 2.0)<br>• Health related quality of life (EQ5D-5L)<br>• Symptoms of breathlessness<br>- at rest and on exertion (Edmonton VAS & Modified MRC Dyspnoea Scale)<br>• Sputum burden (VAS) |

#### Adverse events

Adverse events are defined as any untoward medical occurrence in a patient or clinical investigation subject administered an investigational intervention and any such event does not necessarily have to have a causal relationship with this intervention [45]. Adverse events (AEs) occurring during physiotherapy treatments have been found to occur at a lower rate than those occurring in general intensive care of critically ill patients [39]. It is recognised that the patient population with critical illness will experience a number of common aberrations in laboratory values, signs and symptoms due to the severity of the underlying illness and the impact of standard therapies. These will not necessarily constitute an adverse event unless they are considered to be of concern or directly related to the study or the intervention in the investigator’s clinical judgement. Adverse events will be collected from randomisation to 48 hours post cessation of this protocol only. The occurrence of any adverse events, any impact on the intervention or need for medical review or medical intervention, and the outcome will be collected by research trained staff and recorded in the electronic REDCap case report form (CRF). The patient should be followed until the event is resolved or explained (to day 180 post-randomisation). Frequency of follow-up is left to the discretion of the investigator.

#### Adverse event definition

- Hypotension with MAP < 60, or 5mmHg below the physiological target set by the treating intensivist, for greater than 2 minutes or requiring remedial correction with vasopressors during or within 30 minutes of respiratory physiotherapy
- Hypertension with SBP > 180mmHg for greater than 2 minutes or requiring medical attention during or within 30 minutes of respiratory physiotherapy
- SpO2 < 85% for greater than 2 minutes, during or within 30 minutes of respiratory physiotherapy
- Haemodynamically significant change in heart rhythm (e.g. bradycardia, ventricular ectopy or rapid atrial fibrillation with blood pressure compromise) during or within 30 minutes of respiratory physiotherapy.

#### Serious adverse event definition

- Unplanned extubation during or immediately after respiratory physiotherapy treatment
- Pneumothorax during or 30 minutes following intervention requiring drainage, confirmed by radiologist’s report
- Serious cardiac arrythmia during or within 30 minutes or respiratory physiotherapy e.g. sustained SVT, VT or VF.
- Cardiac arrest during or within 30 minutes of respiratory physiotherapy

#### Reporting of adverse events

All serious adverse events must be reported to the coordinating centre within 72 hours or two working days (whichever occurs first) of the investigators becoming aware of the event. The investigator should notify the Institutional / Ethics Committee of the occurrence of the serious adverse event in accordance with local requirements. A safety analysis will be conducted by the DSMB at 50% recruitment.

#### Data management

Study data will be entered directly into REDCap, a secure web-based CRF. All paper case report forms will be kept in a secure, locked research office within the ICU at each site, and the study database will be kept on a secure, password protected computer. The database will only be accessible to named investigators. A paper screening log will also be kept separately at each study site, in a secure, locked office. This log will only contain the date screened, medical record number, patient initials and age. Enrolled patients will be allocated a unique study number to protect their identity within the research database, and all data will be entered into the CRF under the patient’s study number. A separate enrolment log will be kept at each study site, also in a secure, locked office. The enrolment log will contain the patients name, UMRN, and their study number so that patients may be identified to obtain 90-day follow-up and hospital costing data.

#### Trial monitoring

A detailed study monitoring plan will be prepared by the study management committee prior to commencement of the study. On site source monitoring will be conducted by the co- ordinating centre and will include 100% source data verification for the eligibility criteria and primary end point. All study data will be managed according to Good Clinical Practice and site confidentiality requirements. All data will be kept for 15 years before being destroyed according to local privacy requirements.

#### Data monitoring

Source data verification will occur at all sites for the first 5 participants enrolled and include eligibility, consent and primary outcome data. Subsequent source data verification may be undertaken at the discretion of the trial manager. An independent Data Monitoring Committee (DMC), consisting of experts in intensive care medicine, intensive care physiotherapy, and biostatistics will be established before patient enrolment. No interim analysis is planned, however, all AEs will be provided to the DSC who reserves the right to call for a blinded or unblinded interim analysis at any point in the conduct of the trial.

## Outcomes

This is a hybrid implementation-effectiveness design, therefore implementation (feasibility, acceptability, safety) and effectiveness outcomes will be investigated in parallel and be equally important. The study outcomes for both implementation and effectiveness are outlined in Table 2.

**Table 2:** Implementation and effectiveness outcomes.

| **Implementation Outcomes** |
| --- |
| *Feasibility Outcomes* |
| - Patient recruitability<br>- Patient treatability (treatable traits)<br>- Treatment fidelity of new VHI (Rx arm)<br>- Group separation<br>- Physiotherapy compliance with guideline care<br>- Barriers to implementation<br>- Multidisciplinary staff satisfaction with guideline care may be explored as a sub study using focus groups or questionnaire (potential Honours project). |
| *Safety Outcomes* |
| - Occurrence of predefined adverse events (AE) & serious adverse events (SAE)s<br>- Inflammatory biomarkers (CRP and WBC count) at baseline, 7 days or ICU D/C. |
| **Effectiveness Outcomes** |
| *Primary Outcome* |
| - Disability at day 90 (WHODAS 2.0) |
| *Secondary Outcomes* |
| - Ventilator free days to day 28<br>- Antibiotic free days to day 28<br>- Vasopressor free days to day 28<br>- Discharge destination<br>- Mortality at Day 28 and day 90<br>- Days alive and out of hospital at day 90 |
| - Health-related quality of life at day 90 |
| *Exploratory Outcomes* |
| - Incidence of new pneumonia during the period of invasive mechanical ventilation<br>- Highest mobility at ICU discharge (IMS)<br>- Symptom severity at ICU discharge and hospital discharge<br>- Patient secretion burden (Visual analogue scale 1-10)<br>- Breathlessness (Edmonton VAS, Modified MRC Dyspnoea Scale)<br>- Changes in lung ultrasound scores [46, 47] with new VHI treatment may be explored in a smaller group of patients (10-30) as a substudy at FSH.<br>- Patient & family experience with care may be explored as a future substudy (potential physiotherapy Honours/Masters project). |
| **Cost Analysis** |
| Data will be collected for duration of trial regarding:<br>- Duration of physiotherapy episodes of care in ICU<br>- ICU length of stay<br>- ICU readmission<br>- Respiratory rescue therapies used during ICU stay: re-intubation, bronchoscopy, unplanned emergency respiratory physiotherapy<br>- Surgeries during the hospital stay<br>- Episodes of physiotherapy care (respiratory and rehabilitation) received between ICU and hospital discharge<br>- Hospital length of stay |

## Data analysis

Descriptive statistics will consist of means and standard deviations, or medians and interquartile ranges for continuous data, and frequency distributions for categorical data. Univariate group comparisons will be done using independent t-tests or non-parametric Mann-Whitney U tests for continuous data & Chi-squared or Fisher’s Exact tests for categorical data. Mixed effects generalised linear models, including random effects for patients and site, will be used to examine longitudinal (discharge and 3-months) primary outcome WHODAS 2.0 and secondary outcomes EQ5D-5L and Borg Dyspnoea score. Results will be summarised using estimated mean differences and 95% confidence intervals (CI). Mixed effects logistic regression models will be used to examine longitudinal mortality outcomes, with results summarised using odds ratios (OR) and 95%CI. Ventilator free days will be described using negative binomial models and summarised using estimated means and 95%CIs. Length of stay outcomes will be examined using Kaplan-Meier survival probabilities and described using median (95%CI) days duration, with Log Rank tests used for group comparisons. Cox regression models may be used to derive hazard ratios (HR).

Quality-adjusted life years (QALYS) will be calculated using utility scores from the EQ-5D- 5L collected at 3-months post randomisation. An *a priori* subgroup analysis according to site is planned to investigate any between group difference that is site specific. An interim analysis is planned at 50% recruitment to investigate safety and treatment effect on the primary outcome. Data will be analysed using Stata 18.0 (StataCorp LLC, College Station, TX).

### Sample size

A sample of n=196 (n=98 per group) has 80% power (one-tailed, alpha=0.025) to detect a decrease of 20% MCID from a mean WHODAS score of 26.2 to 20.96, SD=15 (effect size f=0.131), between 2 groups over 2 timepoints in a mixed effects generalised linear model, based on acute respiratory failure sample mean and MCID in Higgins et al [48]. Including a 15% contingency for loss to follow up is n=225 (n=113 per group).

## DISCUSSION

This research aims to evaluate the implementation of a rigorously developed physiotherapy clinical practice guideline for critically ill adults requiring invasive mechanical ventilation for pneumonia, and to determine the effectiveness of adding a set dosage of standardised VHI treatment to standard guideline care on both patient-important and health system outcomes.

It is hypothesised that implementation of physiotherapy guideline care will improve patient outcomes, streamline clinical practice and reduce unwanted clinical practice variability [10, 16] and inefficiency, and will be valuable for training of junior clinical staff to the complex ICU environment.

Implementation outcomes will explore feasibility of guideline implementation, with and without the addition of a standardised set dosage of VHI intervention on: safety; patient recruitability; treatable traits which are amenable to respiratory physiotherapy intervention during the acute intubated period; treatment fidelity of standardised VHI intervention; ability to achieve group separation; physiotherapy compliance with guideline care; barriers to implementation; and multidisciplinary staff satisfaction with guideline care. To the authors’ knowledge, this will be the first study to:

a. explore and document clinical phenotypes of critical pneumonia which are more amenable or less amenable to respiratory physiotherapy intervention.
b. explore safety of physiotherapy guideline care, plus the addition of standardised VHI intervention at a set daily dosage, during the period of critical illness, against pre- defined safety criteria.

Effectiveness outcomes will determine the efficacy of a course of physiotherapy guideline care, with and without the addition of a standardised set dosage of VHI intervention, on patient-important outcomes of disability, ventilator-free days, 90-day mortality, time spent in hospital, symptom severity and health-related quality of life for ICU patients with pneumonia requiring mechanical ventilation, and to determine the cost benefit to the health system. To the authors’ knowledge, this will be the first study to do so.

The results of this study will provide valuable clinical guidance for clinicians regarding the safety of respiratory physiotherapy intervention, the pneumonia phenotypes which are most amenable and responsive to respiratory physiotherapy intervention during the acute period of invasive mechanical ventilation, thus enabling care to be streamlined and clinical inefficiency and waste to be minimised, and will provide essential information regarding the impact of physiotherapy care during the ICU stay on patient-important outcomes for those admitted with critical pneumonia.

## CONCLUSION

Critical pneumonia is a common cause for admission to ICU and the most common cause of ICU admission due to sepsis. Previous research has found that this patient cohort commonly receives respiratory intervention by physiotherapists during the acute period of invasive mechanical ventilation, however physiotherapy care is highly variable. This study will evaluate implementation of a physiotherapy best practice guideline and to address the critical knowledge gaps regarding safety, efficacy, impact on patient-important outcomes and cost-effectiveness.

## Funding

The Coordinating Principal Investigator, Dr Lisa van der Lee, is supported to conduct this study through the award of a Raine Clinician Research Fellowship from the Raine Medical Research Foundation. Prof Anne-Marie Hill is supported by a NHMRC Investigator grant (EL1) and the Royal Perth Hospital Research Foundation.

## Data Availability

This is a protocol paper, there is no data available at present.

## Acknowledgements

The authors sincerely thank the physiotherapy and intensive care staff at participating sites.

## Conflict of Interest

No conflicts of interest have been declared by the authors.

## Notes

### Competing Interest Statement

The authors have declared no competing interest.

### Clinical Trial

ACTRN12624001130550

### Author Declarations

Ethics approval was granted from the South Metropolitan Health Service Human Research Ethics Committee on 24/07/2024. Reference number RGS6734.

